# The Beat Goes On: A Mixed-Methods Analysis in Developing Effective Police Leaver Transitions

**DOI:** 10.64898/2026.03.26.26348236

**Authors:** Eleftheria Vaportzis, Warren Edwards

## Abstract

The end-of-career stage of the police lifecycle represents a profound shift in identity and psychological stability, yet it remains historically neglected in research. This mixed-methods study investigated perspectives of UK police leavers and those approaching retirement (N = 325) regarding desired improvements to organisational support. Content analysis identified four themes: ‘Holistic support and long-term welfare’, ‘Institutional culture and professional worth’, ‘Navigating the structural transition’, and ‘Individual and systemic perspectives’. Findings suggest that the psychological contract between the officer and the organisation is often breached at the exit point, shifting from a relational bond to a transactional disposal. Middle-ranking officers and early leavers report the highest levels of institutional abandonment. To address these gaps, this paper makes recommendations for developing effective transitions. By implementing post□service welfare, and adopting structured resettlement models, police organisations can fulfil their duty of care and mend the psychological contract for those who have served.

---

Policing is widely recognised as one of the most psychologically and physically demanding professions, characterised by chronic exposure to trauma and high-stakes decision-making (Oliver *et al*., 2023). While extensive research has focused on the wellbeing of active officers, the end-of-career stage of the police lifecycle has historically been neglected (Bullock *et al*., 2020a). The transition from a long-term career in the police service to civilian life is not merely a change in employment; it is a profound shift in identity, social standing, and psychological stability (Lennie *et al*., 2025). For many, this change of employment status results in a sudden sense of institutional abandonment, where the organisation’s focus remains on the administration of the exit while neglecting the holistic welfare required to navigate life after the service (Porter and Lee, 2024). The issue is intensified by a growing retention crisis in British policing. According to Home Office (2024), voluntary resignations have hit a record 6.2% leaver rate in England and Wales. This instability is further exacerbated by the McCloud remedy, the legislative response to a 2018 court ruling which found that the 2015 pension reforms were age-discriminatory (Home Office, 2019). The government’s ongoing implementation of this remedy has been marked by technical complexity (HM Revenue & Customs, 2023); for many officers nearing the end of their career, this has created a climate of financial limbo, further complicating the decision-making process of navigating the transition. Current support systems are failing to adapt, remaining geared toward lifelong careers rather than the modern reality. Addressing this transition is not just a matter of individual welfare but a strategic necessity for maintaining institutional integrity during a period of record leaver rates.

This experience of abandonment is best understood through the lens of the Psychological Contract (Rousseau, 1989). Unlike a formal employment contract, the Psychological Contract represents the latent, mutual obligations and unwritten expectations between an officer and the force. In the police family, this contract is deeply relational: officers trade decades of service and trauma exposure for an implicit promise of long-term security and professional respect (Charman & Bennett, 2022). Rousseau (1989) also distinguishes between a breach (the cognitive recognition that expectations have not been met) and a violation (the emotional response that follows). However, as Phythian et al. (2022) suggest, a gap remains in delivering evidence-based, targeted approaches that align these individual employee perspectives with organisational goals. When this alignment fails at the point of exit, it constitutes a contract breach, transforming a lifelong relational bond into a purely transactional termination (Rousseau, 1989). This breach is compounded by the psychological weight of the role; with 45% of officers reporting burnout (Oscar Kilo, 2025), exiting in such a depleted state without structured support often culminates in a profound sense of professional worthlessness.

As UK policing faces urgent challenges regarding officer retention and post-service mental health (Jones *et al*., 2025), understanding these unmet needs is a matter of both institutional urgency and moral duty. By exploring the voices of those who have navigated or currently approaching this transition, this study provides an evidence base for reforming the exit journey and mitigating the emotional exit described by leavers (Lennie et al., 2025). The primary aim of this study was to explore the perspectives of current and former UK police officers, including those who are within 12 months of their transition, have retired or left the Force before retirement, to identify necessary reforms in organisational support throughout the exit process. Specifically, the study investigated participant perspectives on the desired improvements to organisational support mechanisms both prior to and following their departure from the service. By addressing this question, the study seeks to provide a practical evidence base for developing targeted, evidence-based welfare interventions that preserve officer wellbeing and institutional integrity. Ultimately, this work intends to highlight how police forces can honour their unwritten commitments to the workforce, ensuring that the conclusion of a career is marked by professional respect rather than institutional abandonment.

## Method

### Recruitment and survey

‘The Beat Goes On’ was a UK-wide survey conducted between November 2025 and March 2026. Eligibility criteria included being a former police officer in the UK, either retired or having left before retirement, or planning to retire within the next 12 months. The survey was distributed online via Online Surveys (www.onlinesurveys.ac.uk). Data collection continued until two consecutive days passed without any new completions. Duplicate entries were minimised using IP tracking and timestamp checks provided by the Online Surveys platform. Respondents were recruited using multiple outreach methods targeting police officers (e.g., social media groups, police associations). The study was promoted through newsletters, websites, and mailing lists of relevant organisations across the UK [e.g., National Association of Retired Police Officers (NARPO), Police Superintendents’ Association, College of Policing]. These organisations were contacted directly and received study materials including a poster with the survey link for dissemination. Snowball sampling was encouraged, and respondents were invited to share the survey with others who may be eligible. All recruitment materials included eligibility criteria, ethics approval reference, and contact details for the research team.

While the broader survey addressed various aspects of officer wellbeing (Vaportzis and Edwards, under review), the present study focuses specifically on the qualitative responses to the open-ended question: ‘What changes would you like to see in how the police force supports officers before and after leaving the service?’ This question captured broader perspectives on organisational support, transition experiences, and unmet needs. Responses offered personal insights, recommendations, and concerns, highlighting gaps in support and areas for improvement in welfare, career transition, and post□service wellbeing. Ethics approval was granted by the Humanities, Social, and Health Sciences Research Ethics Panel at the University of Bradford (Reference Number E1350). All respondents provided written informed consent.

### Sample

A total of 370 responses were initially collected. However, 45 (12.2%) respondents were excluded from the final analysis as they either left the primary research question blank or indicated they had no specific recommendations to offer. The analytical sample was, therefore, 325 (87.8% of the overall respondents). Respondents included retired officers (n = 251, 77.2%), comprising those who retired on pensionable years, took early retirement, or retired on medical grounds; early leavers (n = 39, 12.0%), comprising those who left from the service prior to reaching retirement age; and soon-to-retire officers (n = 35, 10.8%), comprising active-duty officers planning to leave the service within the following 12 months. The total sample comprised 238 males (73.2%), 86 females (26.5%), with 1 not reporting their sex (0.3%). The mean age was 61.44 (*SD* = 10.43, range 24-93). Most of the respondents identified as White British (n = 312, 96.0%). The majority of the sample served in England or Wales (n = 298, 91.7%), with the remaining representing Scotland (n = 14, 4.3%) and Northern Ireland (n = 13, 4.0%). Within the retired group, the majority completed a standard or full-length service (n = 101, 39.9%). Medically retired officers and those who were fully retired and not working each accounted for 28 (11.1% per group). A further 17 (6.7%) were engaged in policing-related volunteer work, 11 (4.3%) retired early, 17 (5.2%) were working part-time, and 28 (8.6%) reported receiving a pension while remaining engaged in some form of employment. Service characteristics are presented in Table 1.

**Table 1.** Service characteristics of respondents by exit status.

|  | Retired | Early leavers | Soon-to-retire | Total sample | *p* |
| --- | --- | --- | --- | --- | --- |
|  | n = 251 | n = 39 | n = 35 | N = 325 |  |
|  | n (%) | n (%) | n (%) | N (%) |  |
| Highest rank |  |  |  |  | .002*** |
| Constable | 114 (45.4)* | 29 (74.4)*** | 19 (54.3) | 162 (49.8) |  |
| Sergeant | 63 (25.1) | 7 (17.9)*** | 3 (8.6) | 73 (22.5) |  |
| Inspector | 37 (14.7) | 1 (2.6)* | 5 (14.3) | 43 (13.2) |  |
| Chief Inspector | 22 (8.8) | 0 (0.0) | 2 (5.7)*** | 24 (7.4) |  |
| Superintendent | 10 (4.0) | 1 (2.6) | 5 (14.3)** | 16 (4.9) |  |
| Chief Superintendent | 5 (2.0) | 0 (0.0) | 1 (2.9) | 6 (1.8) |  |
| Serving years M(SD, range) | 39.95 (5.30, 14-61) | 26.73 (8.40, 13-50) | 38.60 (4.20, 27-52) | 38.29 (6.99, 13-61) | < .001*** |
| Years since exit M(SD, range) | 23.34 (9.82, 0-51) | 20.35 (12.73, 1-53) | -- | 20.46 (12.05, 0-53) | < .001*** |

\* $p < .05$ , \*\* $p < .01$ , \*\*\* $p < .001$ . $p$ -values reflect results from one-way ANOVA for continuous variables (i.e., serving years) and Chi-square tests for categorical variables (e.g., rank). Missing responses may result in non-rounded values.

### Data analysis

The data were analysed using a deductive content analysis approach (Hsieh and Shannon, 2005). Respondents’ answers were mostly made in short-comment style, although some respondents gave longer answers. Using Excel, the responses were cleaned by the first author (E.V.), correcting spelling and grammatical mistakes, and put in a table. Long reflections were condensed, ensuring all important topics were captured.

The first step of content analysis is to code the data. The codes are then collected under categories before being clustered together to find themes that best describe the dataset. Abiding by this framework, specific examples given by respondents (e.g., I still think about things every day) were assigned a code (e.g., Mental health), followed by a category (e.g., Psychological welfare). The categories were counted by number of respondents and not number of times one participant mentioned a particular category.

The first author coded the first 30 responses. To ensure the trustworthiness and credibility of the findings, the second author (W.E.), who provides lived experience of the police transition, acted as a reflective practitioner during the coding process. The second author assisted in refining category definitions to ensure they captured the cultural nuances of the policing environment, reviewing the coding frame and thematic structure to ensure the interpretations were grounded in the lived reality of the respondents. Discrepancies were discussed until a consensus was reached, resulting in a formal coding template. The first author subsequently coded the remaining data following the template. Upon completion of the first coding phase, the first and second authors discussed appropriate codes for responses that did not fit the coding template. The first author then conducted a final consistency check across all coded data. To ensure reliability, the second author reviewed a random 10% sample of the final coded data to verify the consistent application of the template. By involving a co-author with direct experience of the phenomenon, the deductive categorisation was validated against lived reality, reducing researcher bias and strengthening the ecological validity of the analysis. Consequently, these validated codes were grouped into categories and assigned numerical identifiers. The frequency of comments within these numbered categories was then calculated to identify dominant trends and form overarching themes.

Additional quantitative analyses were performed to explore associations between the four themes and three key demographic variables: current service status, highest rank attained, and voluntariness of exit. These variables were selected as they represent primary determinants of an officer’s relational bond and hierarchical experience within the police service, which are theorised to influence perceptions of contractual fulfilment and the adequacy of institutional support (Rousseau, 1989, Charman and Bennett, 2022). The results were analysed by a series of Multivariate Analysis of Variance (MANOVA) and Covariance (MANCOVA) using SPSS v.28 (IBM, 2021). To ensure sufficient statistical power, rank was categorised into four groups: Constable, Sergeant, inspecting ranks (Inspector and Chief Inspector), and superintending ranks (Superintendent and Chief Superintendent). This grouping was necessitated by the lower frequency of senior-ranking respondents within the sample. Initially, a series of 3 (Service status: Retired, Early leaver, Soon-to-retire) × 4 (Rank: Constable, Sergeant, Inspecting ranks, Superintendent ranks) × 2 (Voluntariness: Voluntary, Involuntary exit) between-subjects MANOVA was conducted. Pairwise differences were explored using Bonferroni post-hoc tests. The analysis was refined into a MANCOVA by including ‘Service years’ as a continuous covariate. Pairwise comparisons for significant effects were conducted using Estimated Marginal Means with Bonferroni adjustment. This approach allowed for the examination of rank and status effects while statistically controlling for exposure in the policing environment. Multivariate significance was determined using Wilks’ Lambda (Λ). Assumption testing revealed violations of the assumption of equality of variances (Levene’s test, *p* < .001) across all themes based on the mean. However, the more robust median-based Levene’s tests remained non-significant (*p* > .05), suggesting that the variance issues were likely driven by the unequal group sizes rather than inherent data instability. To ensure statistical rigour, bootstrapping (1,000 samples) was applied to calculate bias-corrected confidence intervals. Alpha was set at .05.

## Results

### Overview of responses

Many respondents provided detailed, multifaceted accounts; consequently, 218 respondents (66.3%) gave answers that spanned two or more categories. This complexity extended to the thematic level, with 493 thematic occurrences identified across the four overarching themes: ‘Holistic support and long-term welfare’ (n = 200); ‘Institutional culture and professional worth’ (n = 138); ‘Navigating the structural transition’ (n = 119); and ‘Individual and systemic perspectives’ (n = 36). All categories, themes and example quotations reflecting those themes are given in Table 2. Figure 1 illustrates the thematic polarisation of early service leavers, where intense focus on psychological welfare and professional worth (Themes 1 and 2) contrasts with a significant decline in structural transition concerns (Theme 3) compared to those soon-to-retire and retired.

**Figure 1.** Heatmap of thematic frequency by service status

**Table 2.**
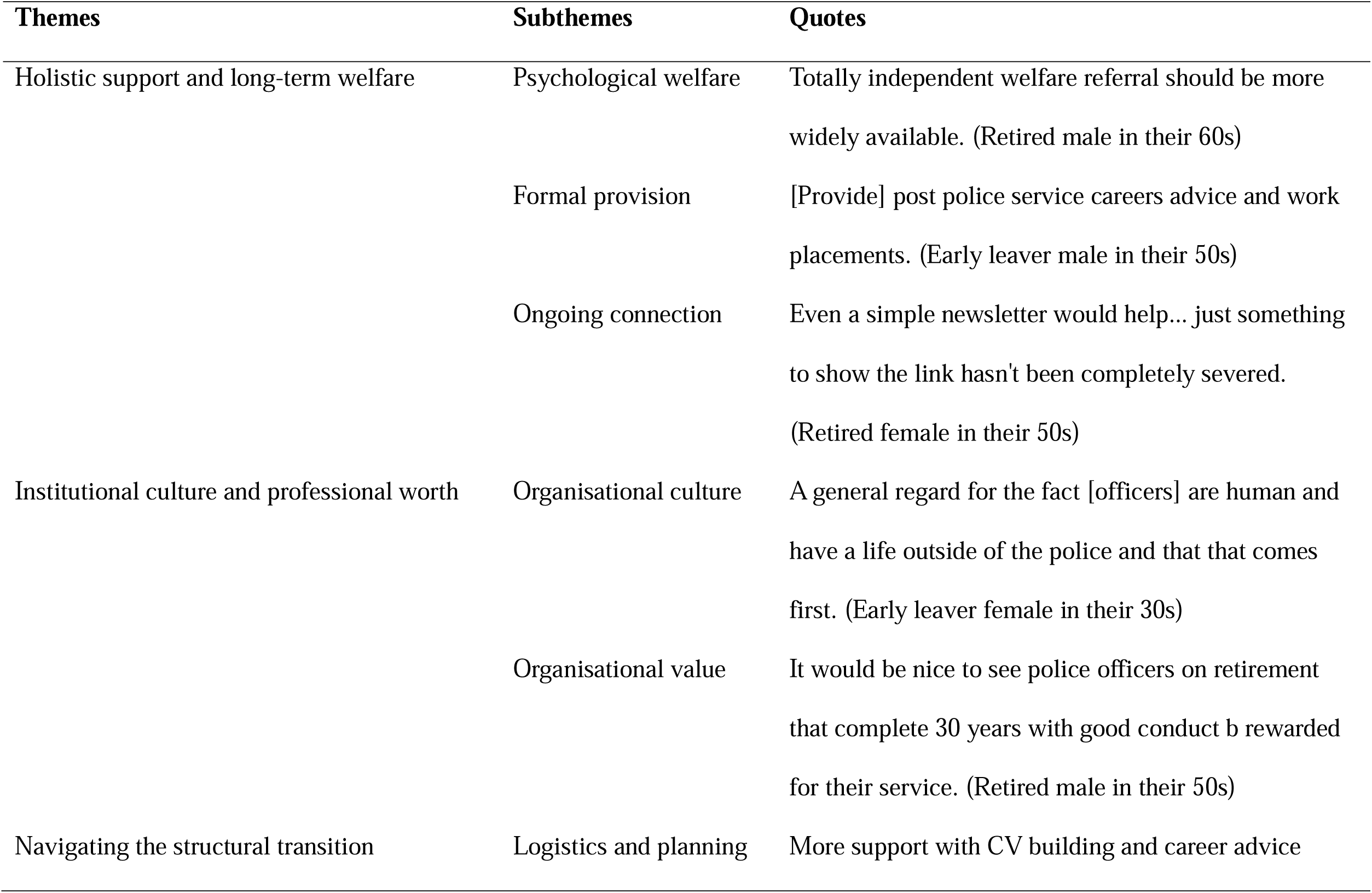
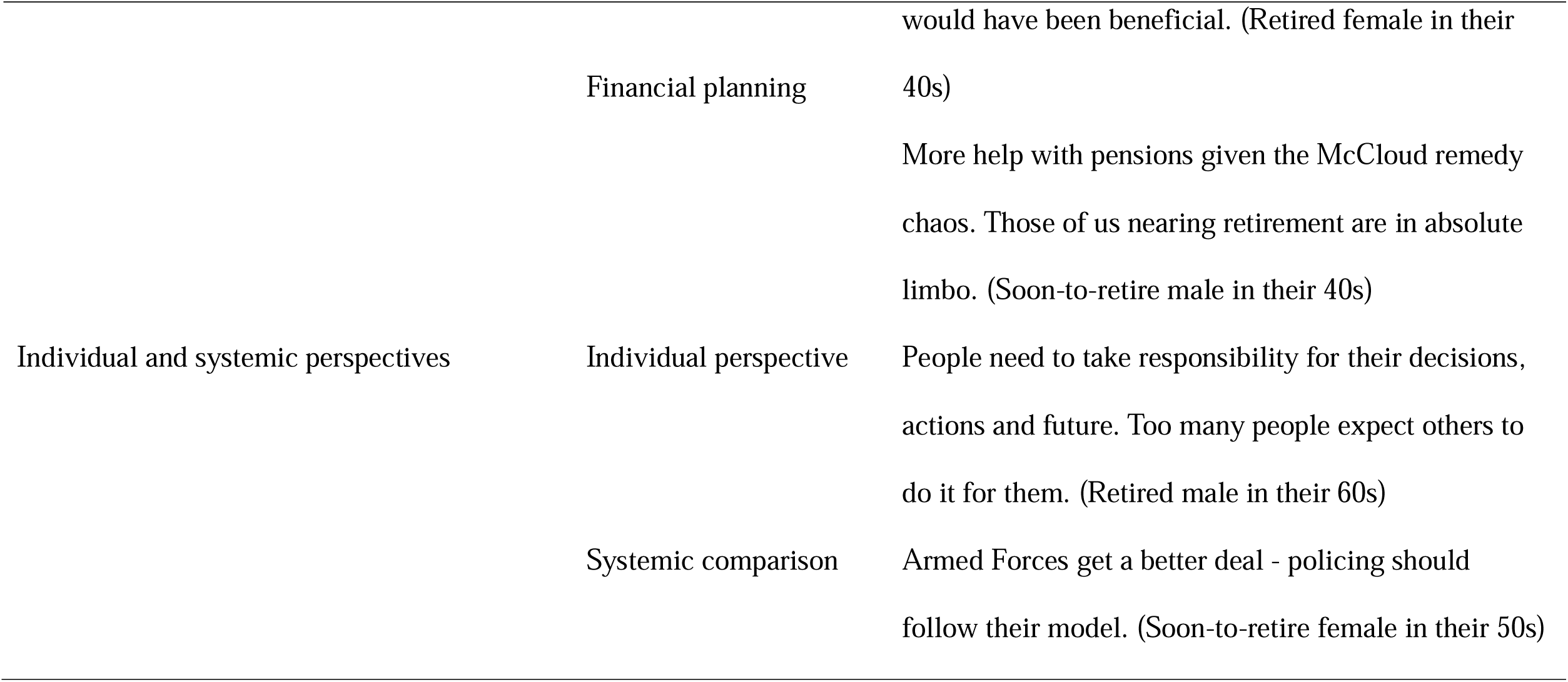
Respondent recommendations for organisational change: a thematic summary with representative quotes.

#### Holistic support and long-term welfare

Respondents expressed a significant need for support that addresses the emotional and psychological complexities of leaving the police service. Three subthemes were developed under this theme: psychological welfare, formal provision, and ongoing connection.

#### Psychological welfare

Respondents emphasised that the transition often triggers underlying psychological issues or a decline in emotional wellbeing. Many noted that the unpleasant experiences they lived during their service did not disappear upon leaving: *I did 28yrs on response and CID [Criminal Investigation Department] with all the unpleasantness that entails. I still think about things every day… in my first six months of retirement I found myself becoming quite upset and dwelling on things quite a lot. (Retired male in their 60s)* Furthermore, there is a recurring sentiment that the Force’s current exit strategy is heavily skewed toward administrative and financial logistics, while the psychological transition of losing one’s professional identity is largely ignored: *I feel that there was a focus on financial needs as opposed to being mentally prepared for stopping work. (Retired male in their 60s)* Respondents highlighted a systemic failure in providing a safety net once they left the Force. There is a common perception that trauma-related issues are treated as an internal employment issue, meaning that once an officer leaves, they lose access to the support systems they need: *Counselling should not be restricted to 6 sessions during service. There is no follow up after leaving the service. (Early leaver male in their 50s)*

#### Formal provision

This subtheme addresses the specific assistance programs and structured support systems offered by the Force. While some respondents acknowledged the existence of retirement seminars and health checks, the prevailing sentiment was that formal provision is currently insufficient for the long-term needs of a departing officer. A significant number of respondents felt that current wellbeing initiatives are superficial. While specific medical or administrative checks were appreciated, they were often viewed as tick-box exercises that failed to address the deeper emotional or psychological complexities of leaving the service: *On a positive [note], wellbeing checks via Bluecrest [were] well received. Everything else simply lip service. (Soon-to-retire female in their 50s)* Respondents frequently described the exit process as a bureaucratic maze. The lack of a centralised, coordinated pathway leaves individuals feeling isolated and responsible for navigating their own transition without adequate professional guidance: *A one stop shop to help you through the process. We are pretty much left to get on with it ourselves. (Soon-to-retire female int their 50s)* Formal provision was criticised for being reactive and strictly tied to active service. Respondents argued that the most effective formal support would be a system that remains accessible after the departure date, particularly for those suffering from service-related health issues: *Greater support is required before retirement, but especially afterwards. (Retired male in their 50s)*

#### Ongoing connection

Respondents highlighted a profound sense of loss regarding the camaraderie and identity associated with the Force. A recurring sentiment was that the professional relationship is terminated too abruptly, leaving many feeling forgotten the moment they depart. Maintaining a relationship with the Force or former colleagues was seen as a vital part of welfare, yet many felt discarded after their final day of service. A primary concern for many respondents was the feeling that the organisation becomes indifferent to an officer the moment they depart. This sudden shift from being a central part of a family to a total stranger can be deeply isolating: *There is a real feeling that as far as the organisation is concerned, you cease to exist on the day you hand in your warrant card. (Retired male in their 60s)* Respondents expressed a strong desire to stay in the loop regarding the Force’s progress and community events. For many, being invited to ceremonial events or receiving updates is a way to maintain the professional identity they built over decades: *Would like to see retired officers kept in the loop for local changes… (Retired male in their 60s) … [and] still being included in things like remembrance for local forces. (Retired female in their 50s)* Ultimately, this theme reveals that welfare is not a finite administrative task, but a long-term requirement that often peaks only after the officer has officially left the service.

### Institutional culture and professional worth

This theme explores the impact of a perceived shift from a traditional police family model to a transactional, business-driven organisational culture. Two subthemes were developed under this theme: organisational culture, and organisational value.

#### Organisational culture

Respondents frequently described a transition from a vocation and a family to a more business-like, data-driven culture where officers feel interchangeable and undervalued. A recurring sentiment was that the Force has moved away from valuing people toward a culture focused on posts and numbers. Respondents noted that this shift leaves long-serving officers feeling disposable once they are no longer operationally useful: *I served for 45 years, 31 as Police Officer and 14 as Police Staff. When I first joined it was a family. Support was there, real comradeship, friendships and good working conditions. By the end of my working life it was a business, faceless, little discipline and you were just a number. (Retired male in their 70s)* Respondents described a culture where the fear of disciplinary action or professional standards overshadows the desire to support officers through difficult operational decisions: *By the end of my career the blame culture was rife with any excuse to put you on discipline through complaints. (Retired female in their 50s)* They also highlighted that the shift toward a business-like environment has fundamentally damaged the social fabric of the Force. This has resulted in the loss of informal peer-support networks that previously helped officers process the psychological demands of the role: *After 30 years in police you become part of the culture and leaving creates a void in your life… (Retired male in their 50s)* The current culture is seen as driving away experienced officers, leading to a drain of knowledge that ultimately harms the service and the public: *[The Force should] positively consider how officers can use skills in police staff/part time/ voluntary role. Acknowledge the expertise the Force has lost. (Retired female in their 50s)*

#### Organisational value

This subtheme centres on how the Force recognises (or fails to recognise) an officer’s contribution. Even when the organisation attempts to show value through formal meetings or awards, the execution often feels like a tick-box exercise, which can leave the officer feeling less valued than if nothing had been done at all: *After 30 years of service I got to spend 10 minutes with the Chief Constable, and given a certificate and plastic force emblem. Did not feel particularly momentous or valued. (Retired male in their 50s)* A recurring sentiment was that an officer’s value is tied strictly to their immediate operational utility. Once an officer nears the end of their career, their decades of experience and dedication are often overlooked: *Near the end of career within the police there is no support and you are thought of yesterday’s person. (Retired male in their 50s)* A key indicator of value for respondents is whether the organisation maintains a connection. The total severance of the relationship upon retirement is interpreted as a sign that the individual was never truly valued as a person, only as a function: *In 23 years after retirement, I have received just one Christmas Card*.

*Pensioners are forgotten. (Retired male in their 70s)* The shift toward a transactional culture has effectively eroded the ‘psychological contract’ between officer and force, leaving many feeling that their decades of service were merely a commodity to be used and then discarded.

### Navigating the structural transition

This theme focuses on the practical, logistical, and financial hurdles of leaving the Force. It highlights a lack of guidance in moving from a highly structured environment to civilian life. Two subthemes were developed under this theme: financial planning, and logistics and planning.

#### Financial planning

Respondents frequently highlighted that financial information is often unavailable or provided too late, making it difficult to plan for life after the Force: *Pre-retirement advice should be provided earlier and over a period of time rather than a 1-day seminar. (Retired female in their 50s)* Retirement is often described as a cliff edge. Officers expressed a desire for a structured wind down period or part-time options to help them adjust to a slower pace of life: *Retirement for me has been a cliff edge. I would have appreciated a part time role after retirement, so I could leave the job gradually, I miss it. (Retired male in their 50s)*

#### Logistics and planning

Many officers mentioned there was no practical help for moving into civilian employment, specifically regarding CV building, career advice, and translating police skills: *When you retire you are given no support whatsoever for your next steps in civilian life. You have no idea what to do unless you have researched things yourself. (Retired male in their 50s)* The analysis uncovered that logistical gaps create distinct structural and psychological challenges, particularly for senior leaders: *[Provide] courses and preparation for life after policing particularly for senior leaders who face a significant downgrade in their ‘perceived net worth’ when they retire. (Soon-to-retire male in their 50s)* This statement reflects how the loss of status and organisational value is experienced at the point of retirement. It suggests that for senior leaders, retirement involves not only logistical adjustment but a profound readjustment of identity as their organisational status and professional standing fall away. Consequently, the structural transition is currently perceived as a cold administrative exit rather than a supported resettlement period, which significantly exacerbates the stress of leaving.

### Individual and systemic perspectives

This theme explores the tension between personal agency and institutional responsibility, often through direct comparisons between the police service and other public sectors such as the military or the NHS. Two subthemes were developed under this theme: individual perspective and systemic comparison.

#### Individual perspective

There is a subset of respondents who believe the onus for a successful transition lies with the individual, or who question whether it is even achievable for the organisation to support everyone: *I feel the individual needs to take more personal responsibility for their own retirement plans (Retired female in their 60s)* This sentiment often extends into a pragmatic scepticism regarding the Force’s role, with some questioning whether universal organisational support is a realistic expectation: *I suppose my question is why should they or is there a requirement to? Perhaps in certain circumstances but I would suggest to try and do this for all is unachievable. (Soon-to-retire male in their 50s)*

#### Systemic comparison

Respondents mentioned the police service lacks the structured resettlement, retraining, and glidepath that are standard in the military: *The military provide courses and practical advice about your next steps including coaching and training, the Police provide nothing. (Retired male in their 50s)* This sense of institutional stagnation is further reinforced by comparisons to the private sector, where even proactive attempts by staff to mirror corporate off-boarding standards are met with organisational inertia: *My [spouse] worked for [a private company]. A year before retirement they were given a folder telling them exactly what [the employer] and what the employee needed/would do in the months up to and beyond retirement. I suggested something similar to [senior management]. They agreed that it was an excellent idea and would forward to the Human Resources Department. Needless to say, nothing changed. (Early leaver male in their 60s)* By contrasting the police service with the military, respondents highlight a systemic failure to provide a professional off-boarding standard. However, the presence of views regarding personal responsibility highlights an internal tension: while many demand better institutional support, others suggest that the current culture makes self-reliance a necessity for a successful transition.

### Quantitative analysis

A 3 × 4 × 3 between-subjects MANOVA was initially conducted to examine the effects of service status, rank, and voluntariness on the four themes. Using Wilks’Λ, a significant multivariate main effect was found for current status (Λ = .949, *F*(8, 736688) = 2430.89, *p* < .001, η^2^ = .02), rank (Λ = .975, *F*(12, 974546) = 796.09, *p* < .001, η^2^ = .01) and voluntariness (Λ = .986, *F*(8, 736688) = 654.49, *p* < .001, η^2^ = .01). Significant interactions were found between current status and rank (Λ = .902, *F*(20, 1221659) = 1924.01, *p* < .001, η^2^ = .02) and current status, rank, and voluntariness (Λ = .982, *F*(4, 368344) = 1641.64, *p* < .001, η^2^ = .018), suggesting that the combined effect of rank and status is influenced by whether an officer’s exit was voluntary or involuntary. Bonferroni post-hoc tests revealed significant differences across all themes between serving, retired, and early-leaving officers (*p* < .001), all ranks (p < .001) and voluntary and involuntary exit types (*p* < .001).

To account for differences in time in service, a MANCOVA with service years as a continuous covariate was run. Although the covariate was not significant (Λ = .985, *p* = .27), it altered the significance of the main effects. The MANCOVA main effects for current status (*p* = .13), rank (*p* = .76), and voluntariness (*p* = .84) were no longer significant. However, the significant interaction between current status and rank persisted (Λ = .907, *F*(20, 1131.9) = 1.68, *p* = .03, η^2^ = .02; see Table 3).

**Table 3.** MANOVA and MANCOVA for the effects of current status, rank, and voluntariness.

| Effect/Interaction | Model | Wilks' $\Lambda$ | $F$ | $p$ | $\eta^2$ |
| --- | --- | --- | --- | --- | --- |
| Current status | MANOVA | .949 | 2430.89 | < .001*** | .03 |
|  | MANCOVA | .964 | 1.57 | .13 | .02 |
| Voluntariness | MANOVA | .987 | 614.36 | < .001*** | .01 |
|  | MANCOVA | .988 | 0.52 | .84 | .01 |
| Rank | MANOVA | .975 | 796.09 | < .001*** | .01 |
|  | MANCOVA | .976 | 0.69 | .76 | .01 |
| Current status × Rank | MANOVA | .902 | 1924.01 | < .001*** | .02 |
|  | MANCOVA | .907 | 1.68 | .03 | .02 |
| Voluntariness × Rank | MANOVA | .941 | 1403.84 | < .001*** | .01 |
|  | MANCOVA | .947 | 1.17 | .29 | .01 |
| Current status ×<br>Voluntariness × Rank | MANOVA | .982 | 1641.64 | < .001*** | .02 |
|  | MANCOVA | 0.985 | 1.31 | .27 | .01 |
| Service years (Covariate) | MANCOVA | 0.985 | 1.30 | .27 | .01 |

\* $p < .05$ , \*\* $p < .01$ , \*\*\* $p < .001$ .

Pairwise comparisons suggested that for ‘Holistic support and psychological welfare’ (Theme 1), a significant difference was observed within the constable rank: those who left the Force early reported a significantly different experience of psychological welfare compared to those who had reached full retirement (*p* = .04). For ‘Institutional culture and professional worth’ (Theme 2), significant differences were found within the sergeant rank: early leavers experience the most significant cultural disconnect. Compared to those currently serving (*p* = .01) and those who had reached retirement (*p* = .01), early leavers reported a more intense sense of being treated as a transactional commodity rather than a valued member of the family. For ‘Navigating the structural transition’ (Theme 3), a significant shift was found within the inspector ranks. The transition into retirement appears to trigger a notably sharper severance of the institutional bond compared to those still serving (*p* = .01). This suggests that for inspectors, retirement represents a loss of belongingness and organisational connection that is not as acutely felt in other ranks. Finally, for ‘Individual and systemic perspectives’ (Theme 4), within the superintendent ranks, a significant difference emerged between those currently serving and those who are retired (*p* = .03). Overall, these results suggest that the retirement cliff-edge is not a uniform experience. The combination of rank and current status serves as a primary driver for the thematic shifts observed, particularly for middle-to-senior leadership ranks.

## Discussion

The transition from a policing career is often seen as an administrative or financial process. However, this study’s findings demonstrate that for many UK officers, this retirement cliff-edge marks a significant breakdown in the relational bond, transforming a career-long commitment into a transactional exit. This transformation is indicative of the New Public Management (NPM) era in policing, where traditional values have been superseded by a focus on metrics, efficiency, and performance indicators (Charman, 2017). This shift redefines the officer not as a lifelong member of a professional community, but as a unit of labor within a transactional system that ends abruptly at the point of exit (Hoggett *et al*., 2019).

By integrating qualitative themes with quantitative analysis, we sought to identify how institutional support can be improved to meet the unmet needs of those leaving the service. The results show that the transition is not a uniform experience, but a challenging psychological restructuring of the self that varies depending on an officer’s rank and status when they leave the Force. This complexity is reflected in our primary quantitative finding. While previous research has often focused on length of service as the driver of transition difficulty (Bullock *et al*., 2020a), our results demonstrate that once service years are controlled for, the dominant factor shaping the transition experience is the interaction between current status and rank. This suggests that the psychological impact of the exit is mediated by one’s hierarchical identity rather than the years served.

Policing is rarely just a job; it is an identity. Our data show that this identity is experienced hierarchically. We found a significant role conflict among sergeants who leave the service early. As the primary bridge between strategic objectives and tactical delivery (College of Policing, 2025), sergeants often tie their professional self-worth to an organisational value that feels unfinished if they exit early. For this group, the cliff-edge is characterised by a more pronounced sense of lost purpose and a professional void compared to other ranks.

The theme of ‘Navigating the structural transition’ is most evident in the experiences of the inspector ranks, where the data reveals a marked transition in their sense of professional belonging compared with the other groups. While serving, inspectors report a strong sense of organisational connection, which diminishes abruptly upon retirement. This suggests that the inspector ranks represent a threshold of organisational integration. For this group, retirement is not simply a change in employment status but a marked severance of a relational bond. This severance necessitates significant identity work; the internal process through which individuals strive to sustain a coherent and valued sense of self (Brown, 2022).

For the most senior ranks, the theme of ‘Individual and systemic perspectives’ highlights a distinctive psychological trajectory. Among superintendents, shifts in perspective emerge only after retirement, reflecting the significant difference observed between serving and retired officers. This marks a transition from holding a position of high organisational value to the relative anonymity of civilian life. While lower ranks maintain a more stable outlook across the transition, senior officers undergo a profound adjustment in their individual perspective once the status and recognition associated with their rank are gone. This adjustment mirrors findings by (Bullock *et al*., 2020b) who argue that for senior officers, the loss of a social identity rooted in authority can lead to a profound sense of injustice if the exit is not handled with dignity.

Perhaps the most striking finding is the universality of these experiences across demographic lines. The lack of significant differences based on sex or exit voluntariness points toward a uniformity effect within the police culture. Whether an officer chooses to leave or is forced out, the psychological impact of leaving the police family remains a dominant, shared experience that transcends sex or the reasons for the departure. While a uniformity effect exists regarding the emotional impact of leaving the family, Figure 1 shows a clear polarisation in priorities. Early leavers are primarily driven by concerns around ‘Holistic support and welfare’, although issues related to ‘Institutional culture and professional worth’ also feature strongly in their experiences. Those approaching retirement are more preoccupied with the ‘Structural transition’ and the uncertainty created by the McCloud remedy (Home Office, 2019). Ultimately, the transition experience is a multifaceted struggle where institutional culture (Theme 2) creates the identity crisis, but a lack of holistic support (Theme 1) and poor structural navigation (Theme 3) prevent a successful resolution. While some officers maintain an individual perspective (Theme 4) that stresses personal responsibility, the majority of respondents across all ranks point toward a systemic failure to honour the psychological contract through the point of exit.

These findings can be understood through the lens of Rousseau’s (1989) Psychological Contract, particularly the distinction between relational and transactional contracts. Officers enter the service with an implicit understanding of mutual loyalty. While the officers in this study fulfilled their side of this contract through decades of service, many perceived a breach at the point of exit. The withdrawal of organisational support and professional recognition that accompanies retirement is experienced by many as a betrayal of this long-standing agreement. This sense of betrayal has a direct effect that hinders future recruitment. As Bullock *et al*. (2020b) note, the police identity is a social identity; when it is damaged at the point of exit, leavers are less likely to act as advocates for the profession. In an era where potential recruits are heavily influenced by the word-of-mouth reputation of the service, disgruntled leavers who feel abandoned by the organisation become a reputational deterrent, making it increasingly difficult to fill the gaps left by the ongoing retention crisis (PFEW, 2024).

This perceived betrayal has significant implications for recruitment. Recent research into police employment branding indicates that the shared lived experiences of leavers, often disseminated through online company reviews and word-of-mouth, act as a deterrent to potential candidates (Miles-Johnson, 2026). If officers exit feeling abandoned (as many of the 325 participants here reported) their negative narratives can discourage the very recruits the Police Uplift Programme was designed to attract (NPCC, 2024, PFEW, 2024). This theoretical progression, and the moderating influence of rank on how officers experience it, is illustrated in Figure 2.

**Figure 2:** The theoretical framework of the police retirement cliff-edge

### Recommendations

Based on the findings, four recommendations are proposed to improve the transition experience for officers:

1. Welfare provision beyond retirement. To prevent officers from falling through the welfare gap once internal support networks are severed, the Force could implement a proactive welfare check system. This would involve structured outreach at 3, 6, and 12-month intervals post-retirement delivered via phone call or the individual’s preferred contact method. This personalises the support process and ensures consistent engagement during the most critical phases of the transition. Furthermore, we recommend the introduction of standardised transition packs to provide a comprehensive roadmap to administrative guidance and direct signposting to external partners such as Police Care UK, ensuring long-term support remains accessible and clear.
2. Voluntary post-service mentorship. To mitigate feelings of institutional abandonment and address the perceived breach of the psychological contract, the Force could establish a voluntary post□retirement mentorship programme that invites retired officers to mentor colleagues within 2-3 years from their own transition. This initiative would reaffirm the organisation’s recognition of a retiree’s career-long contribution, sustaining their sense of professional worth and continued belonging. In addition, active-duty officers would benefit from the transfer of experience, knowledge, and cultural insight, creating a reciprocal relationship that strengthens the individual’s transition.
3. Mandatory pre-retirement transition framework. To ensure equitable support across the service, the Force could implement mandatory attendance at a formal pre-retirement course, which must be completed no later than 12 months prior to an officer’s expected retirement date. This timeline allows sufficient lead time for officers to navigate complex administrative issue, and to begin the psychological adjustment to civilian life. Importantly, this mandate must explicitly include provisions for officers who are on long-term sick leave prior to retirement, ensuring that physical or mental health challenges do not result in a loss of access to vital transition resources. By standardising this requirement, the Force moves away from a postcode lottery of support, establishing a guaranteed pathway to retirement that prioritises the wellbeing of all personnel regardless of their duty status.
4. Mandatory standardised exit interview: To ensure consistency and accountability, the Force could implement mandatory, standardised exit interviews between retiring officers and their line managers (or another senior officer if requested by the interviewee). These sessions must move beyond basic administration to provide a dedicated space for officers to address personal reflections and service-related issues. Furthermore, these consultations should serve as a formal checkpoint to ensure the individual has received their retirement transition pack and has been allocated the necessary time to attend the pre-retirement course. By formalising this final interaction, the Force can actively repair the psychological contract, ensuring that every officer’s departure is marked by meaningful recognition and a verified pathway to post-service support.

### Strengths, limitations, and future directions

The study has several strengths. Firstly, it used a mixed□methods design, which integrates qualitative with quantitative analysis to capture both the structural patterns and the lived psychological realities of the policing transition. By examining the interaction between rank and current status, the research provides a novel contribution to the literature, reframing the retirement cliff□edge as a hierarchical process. The application of Rousseau’s Psychological Contract further strengthens the theoretical grounding, illuminating how perceived breaches and violations shape officers’ emotional responses at exit. However, the study is limited by its cross□sectional design, which restricts causal inference and prevents observation of how identity and wellbeing evolve over time. Self□selection and self□report biases may also influence the findings, and the findings may limit generalisability across UK policing. Future research would benefit from longitudinal designs that track officers before, during, and after retirement; comparative studies across forces with different organisational cultures; and deeper exploration of how psychological contract dynamics vary across career stages. There is also scope to examine how organisational interventions, such as structured transition programmes or rank□specific support, might mitigate the sense of breach and identity disruption identified in this study.

## Conclusion

The transition out of policing represents a profound psychological, professional, and identity shift that extends beyond administrative retirement processes. This study demonstrates that officers’ experiences of this transition are shaped not only by individual circumstances but by the interaction between rank and current status, which influences how the psychological contract is perceived at the point of exit. The findings highlight significant gaps in welfare provision, leadership practice, and structural support, alongside a strong desire among officers for continued connection and recognition. By integrating qualitative insights with quantitative evidence, this study provides a foundation for developing more responsive, rank□sensitive, and psychologically informed transition pathways. Strengthening post□service welfare, and adopting structured resettlement models can help ensure that officers leave the service with dignity, continuity, and a sustained sense of professional worth. Addressing these needs is essential not only for individual wellbeing but for the long□term health and integrity of the policing profession.

## Data availability statement

Data are available from the authors upon reasonable request.

## Biographical sketches

Dr Eleftheria Vaportzis is an Associate Professor of Psychology at the University of Bradford. Her research focuses on survey methodology and behavioural interventions designed to improve mental wellbeing, quality of life, and cognitive functioning across the lifespan. She has extensive experience leading mixed□methods studies and developing evidence□based approaches to support wellbeing in applied settings. She is a Chartered Psychologist with the British Psychological Society and a Fellow of the Higher Education Academy.

Warren Edwards served in the police for 30 years, retiring from Durham Constabulary in 2020 at the rank of Inspector. His extensive career across the service provided him with deep operational and leadership experience, as well as a first-hand understanding of the unique pressures faced by officers. Since retiring, Edwards has acted as a Lived Experience Expert, ensuring that research into policing remains grounded, practical, and impactful.

